# Imposter Phenomenon in licensed physical therapists: prevalence, predictors, and impact

**DOI:** 10.1101/2023.06.27.23291905

**Authors:** Alexandra R. Anderson, Jamie LaPenna, Dustin Willis, Alison H. Chang

## Abstract

**Objective:** Determine the prevalence and predictors of Imposter Phenomenon (IP) among licensed physical therapists (PTs) in the United States.

**Methods:** This cross-sectional observational study utilized an online survey to assess the levels of IP, emotional exhaustion, and job satisfaction among licensed PTs in the United States and collect their professional and demographic information. Participants were recruited from various physical therapy specialty groups representing a range of professional memberships and certifications. A multivariable logistic regression model examined factors associated with IP presence.

**Results:** The study sample consisted of 514 PTs [women (66.1%), ≤ 40 years of age (73.2%), White (77.0%)]. The mean IP score was 60.3 (SD: 15.1, range: 19 to 95). 55 respondents (10.7%) had low IP, 196 (38.1%) moderate IP, 215 (41.8%) frequent IP, and 48 (9.3%) intense IP. The prevalence of IP, defined as frequent or intense IP, was 51.2% in our study sample. Having Manager/supervisor experience (OR=0.55, 95% CI=0.34-0.90) was associated with a reduced odds of IP presence. Holding a bachelor’s or master’s degree (vs. Doctor of Physical Therapy (DPT) degree; OR=2.31, 95% CI=1.07-5.00), a history of or current mental health diagnosis (OR=2.77, 95% CI=1.69-4.54), and emotional exhaustion (moderate vs. low: OR=5.37, 95% CI=2.11-13.69; high vs. low: OR=14.13, 95%CI=5.56-35.89) were each associated with an increased odds of IP presence.

**Conclusions:** IP is highly prevalent among licensed PTs, with more than half reporting frequent or intense IP. Seasoned clinicians with managerial roles seemed to be less susceptible to IP, whereas those with mental health diagnoses, emotional exhaustion, and those without a DPT degree may be more likely to experience IP.

**Impact Statement:** Given the high prevalence of IP and its potential negative impact on burnout and career advancement, it is crucial to increase IP awareness and provide education on strategies and resources to manage it for licensed PTs.

## INTRODUCTION

Imposter Phenomenon (IP), or perceived fraudulence, describes an ongoing fear of exposure as a fraud or imposter, despite objective successes and accomplishments^1^. Those with IP tend to attribute their achievements to luck or good timing and question their professional legitimacy and expertise.

Recent studies have reported high prevalence of IP among physicians and academic faculty, which could have a detrimental effect on psychological well-being and contribute to burnout, ultimately stunting both personal and professional growth^1–3^. A 2020 systematic review outlined the prevalence, predictors, and potential treatment of this long-recognized phenomenon that impacts approximately 70% of working professionals at some point in their career^1^. IP was strongly correlated with depression and anxiety, poor work performance, job dissatisfaction, and burnout, particularly among ethnic minority groups. Their findings suggested those affected, specifically healthcare providers, may be limited in achieving their full professional potential and are more likely to experience burnout.

Professional burnout and mental health comorbidities are on a steep rise, further fueled by the recent Covid-19 pandemic^4^. While job burnout among physical therapists (PTs) is associated with emotional exhaustion and a decreased sense of personal achievement^5^, no studies have examined the prevalence of IP in licensed PTs. The objectives of the present study were to 1) examine the prevalence of IP among licensed PTs, 2) determine demographic and professional factors associated with IP, and 3) examine its relationship with burnout and job satisfaction.

## METHODS

### Study Design and Setting

An observational study using a cross-sectional survey design was utilized. The survey was delivered in English and hosted on Google Forms, a web-based survey tool. Data collection took place from June 10 to July 10, 2022. The study was approved by West Coast University Institutional Research Board and followed the STROBE guidelines.

### Participants, Recruitment, and Consent

Eligible participants were licensed PTs in the United States. A recruitment flyer was shared with peers/colleagues, posted on social media, and distributed from professional organizations including Academies of the American Physical Therapy Association (APTA) that allow electronic survey distribution to their members, including orthopedics, neurological, geriatrics, home health, and aquatics. Members of the American Academy of Orthopaedic Manual Physical Therapists and both the Illinois Physical Therapy Association as well as the California Physical Therapy Association were also invited to engage in the survey. Each participant provided informed consent prior to taking the survey and was instructed to only complete the survey once.

### Online Survey

The survey was created by the study team of four PTs, each with greater than 5 years of experience in clinical practice. The authors recruited five practicing PTs from various practice settings to pilot test the survey draft. Their feedback was used to refine the survey prior to its distribution. The final survey consisted of 18 demographic and professional questions and 3 sub-surveys to measure the degree of IP, emotional exhaustion/burnout, and job satisfaction.

Demographic questions included age, race, gender, sexuality, years of experience, length at current position, practice setting, practice specialty, terminal degree, credentials (e.g., Orthopedic Certified Specialist, Fellow of the American Academy of Orthopaedic Manual Physical Therapists, etc.), APTA membership, geographical location of practice, manager/supervisor experience, time per week spent on direct patient care, duration of initial evaluations, duration of return visits, and mental health diagnosis. In questions where respondents selected “other” in place of the listed options, a free-text box was provided for more detailed responses.

The 20-item Clance Imposter Phenomenon Scale was used to assess the degree of IP. It is the most widely used instrument known for its good reliability and validity, and ability to distinguish from self-esteem, social anxiety, and self-monitoring^6–8^. The scale uses a 5-point Likert scale for each question with a total score ranging from 20 to 100. A score under 40 is considered low, 41-60 is moderate, 61-80 is frequent, and greater than 80 is intense^9^.

Emotional exhaustion is the greatest predictor of burnout^10^. To assess this key attribute, the survey utilized a 9-item emotional exhaustion scale, a subscale within the Maslach Burnout Inventory^11^. The items were answered on a 5-point frequency scale ranging from 1 (never) to 5 (always) with a total score ranging from 9 to 45^12, 13^. A score of 27 or higher indicates extreme emotional exhaustion level, 19-26 moderate, and under 18 low^13^.

The Job Satisfaction Subscale of the Michigan Organizational Assessment Questionnaire was used to measure job satisfaction, which includes 3 questions on a 6-point Likert scale^14^. The first question was negatively worded, therefore was scored in reverse to align with the remaining two positively worded questions. The final scores range from 3 to 18. Scores were dichotomized into yes (score ≥ 12) versus no (score < 12) job satisfaction. It took approximately 5-10 minutes to complete the survey and there was no financial compensation awarded to participants.

### Data Cleaning

Respondents who were not licensed PTs, such as physical therapy assistants and students, were excluded. One participant did not answer the demographic questions, therefore was also excluded.

### Data Synthesis and Analysis

Following recommended guidelines^9, 11, 14^, Clance IP raw scores were converted to IP levels of low (≤ 40), moderate (41-60), frequent (61-80), and intense (> 80); emotional exhaustion scores to low (<18), moderate (19-26), and high (≥27); job satisfaction scores to yes (≥ 12) and no (<12). Descriptive statistics summarized the characteristics for the overall sample and by IP levels. Missing data (i.e., the response was “blank”) were not imputed.

To assess univariate associations between each factor and IP levels, Chi-Squared tests were conducted. Factors with a P value ≤ 0.20 in the univariate comparisons were considered for inclusion in the final multivariable logistic regression model. For ease of interpretation, IP levels were condensed to a binary outcome of no (combining low and moderate IP) versus yes (combining frequent and intense IP). Multicollinearity check was performed.

Purposeful model selection^15^ was used to produce the final regression model. Briefly, the iterative process proposed in the purposeful model selection recommends (1) removing predictors with a P value ≤ 0.1; (2) adding back a predictor if removing it resulting in ≥ 20% change in any of the remaining predictor’s beta coefficients, because the deleted predictor has provided important adjustment of the effect of the remaining predictors; (3) selecting the model with the highest R^2^; and (4) including predictors that are clinically meaningful/important based on content expertise and prior literature. The Hosmer-Lemeshow goodness-of-fit test checked the fit of the final model. Results were reported as odds ratios (ORs) and 95% confidence intervals (CIs). ORs > 1.0 indicated increased odds and < 1.0 indicated decreased odds. All analyses were performed using SPSS 28.0.1.1.

## RESULTS

A total of 520 survey responses were recorded. After excluding responses from physical therapy students and/or physical therapy assistants (n = 5) and the response without any demographic data (n = 1), our final analysis sample consisted of 514 records. Table 1 summarizes the demographic and professional characteristics of the study respondents. The majority of respondents were women (66.1%), at or under 40 years of age (73.2%), and White race (77.0%). The mean IP score was 60.3 (SD: 15.1, range: 19 to 95). As shown in Figure 1, 55 (10.7%) had low IP, 196 (38.1%) had moderate IP, 215 (41.8%) had frequent IP, and 48 (9.3%) had intense IP. When dichotomized, IP was absent in 251 (48.8%) and present in 263 (51.2%) survey respondents. A sensitivity analysis to compare 4-level vs. 2-level IP categories showed immaterial differences in factors with a P-value ≤ 0.20 (see Supplemental Table 1).

**Figure 1.**
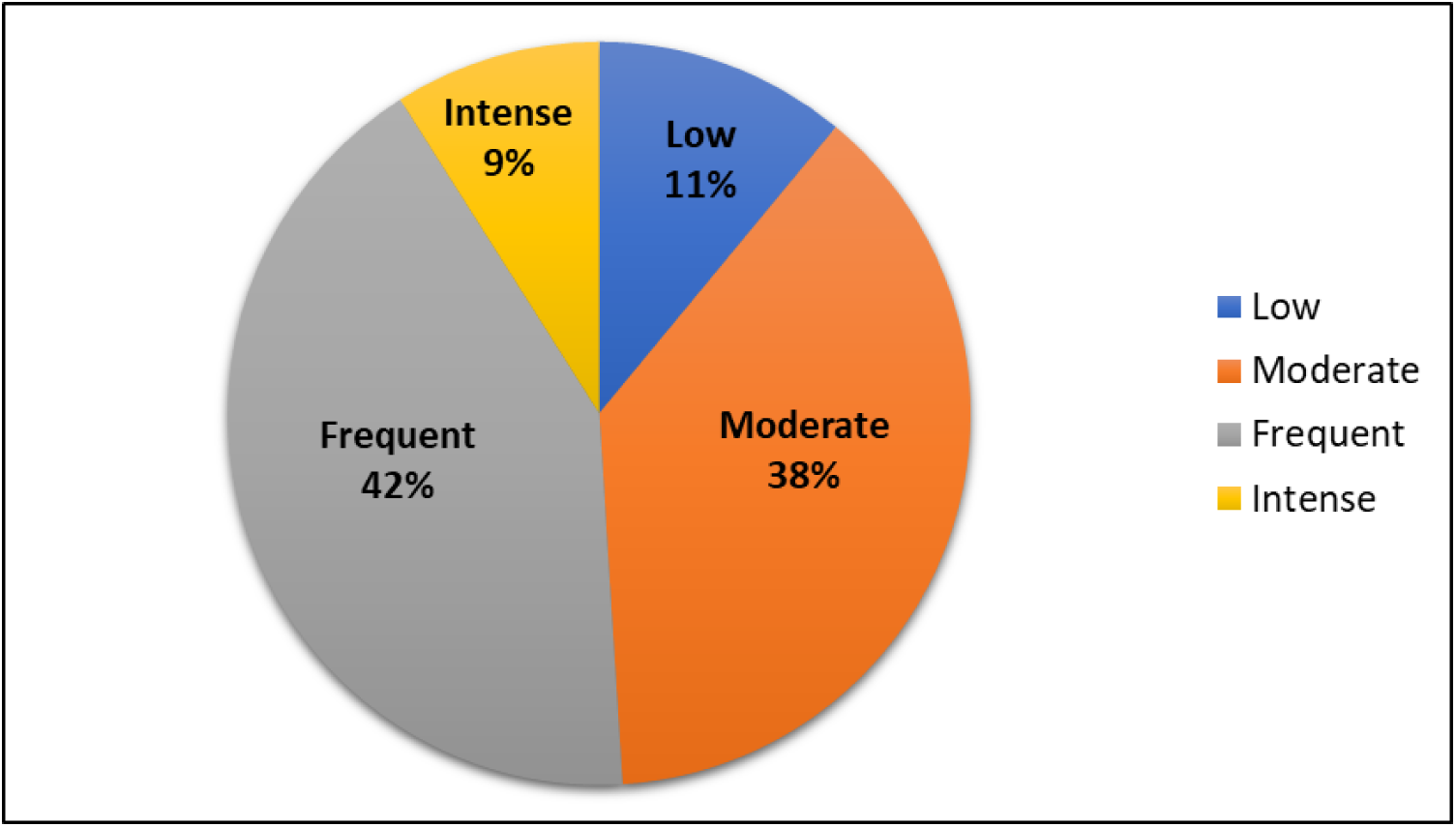
Imposter Phenomenon (IP) prevalence among US physical therapists.

**TABLE 1.**
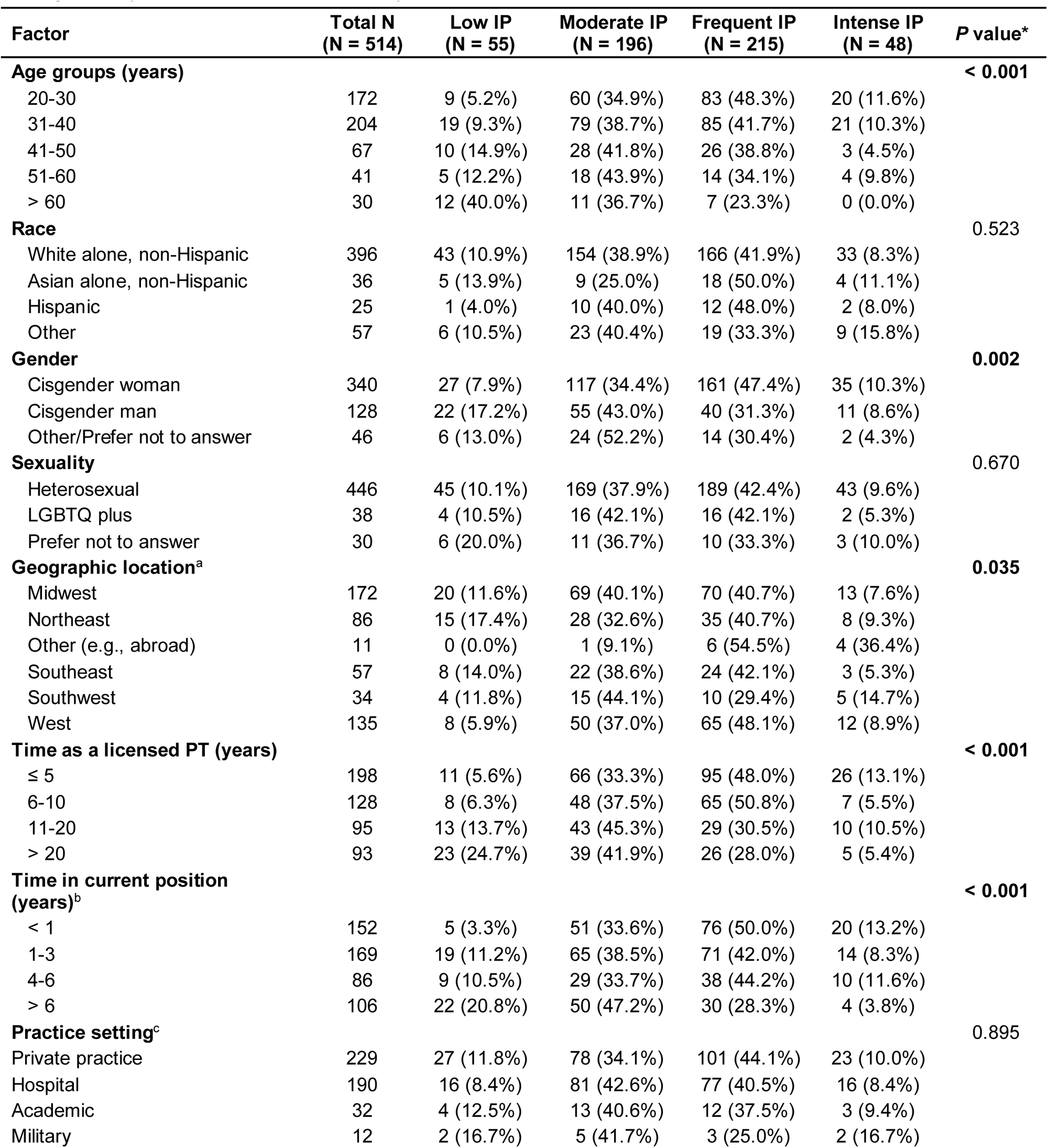
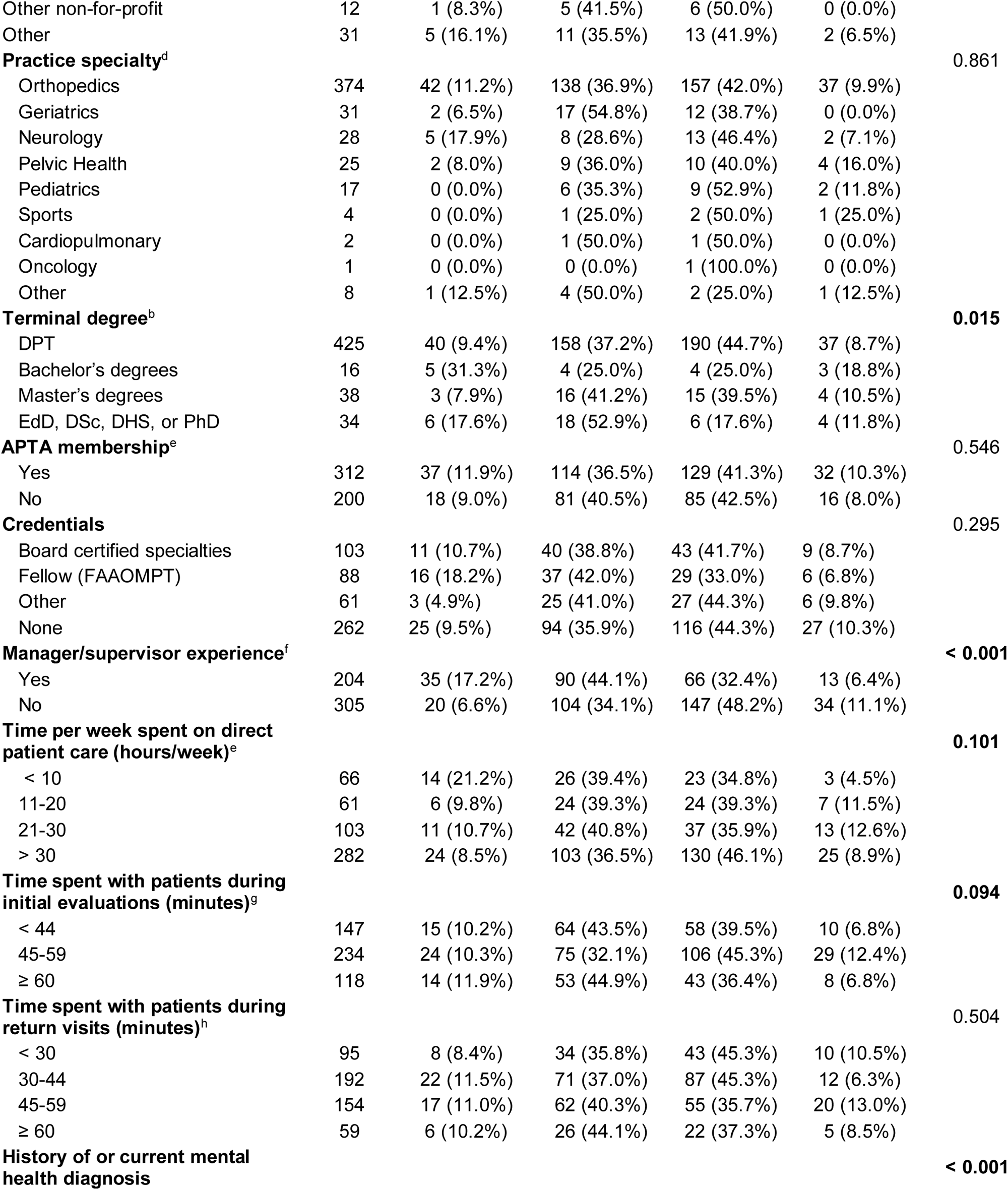
Descriptive Summary of Demographic and Professional Factors related to 4 IP Categories (Univariate Comparisons)

Age was strongly correlated with years of experience as a PT (R = 0.85), therefore was excluded from the model to avoid multicollinearity. The initial logistic regression model included the following 11 predictors: gender, geographic location, time as a licensed PT, time in current position, terminal degree, manager/supervisor experience, time spent on direct patient care, time spent with patients during initial evaluations, history of or current mental health diagnosis, emotional exhaustion, and job satisfaction. Following the principles for purposeful model selection (outlined in the Methods section), we identified the final most parsimonious logistic regression model, shown in Table 2. The Hosmer-Lemeshow goodness-of-fit test had a P value of 0.897, suggesting that there was no significant difference between observed and predicted values.

**TABLE 2.**
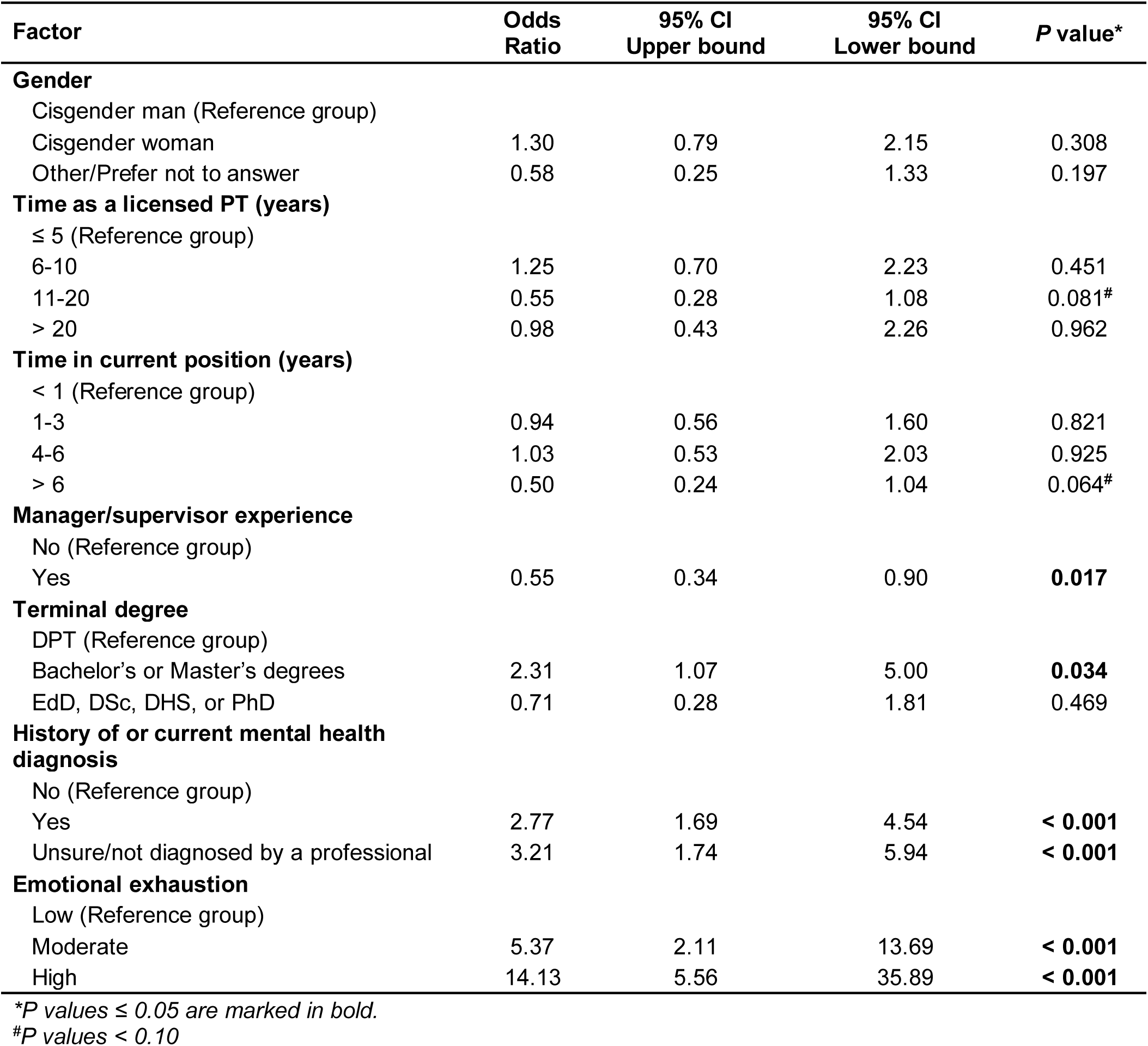
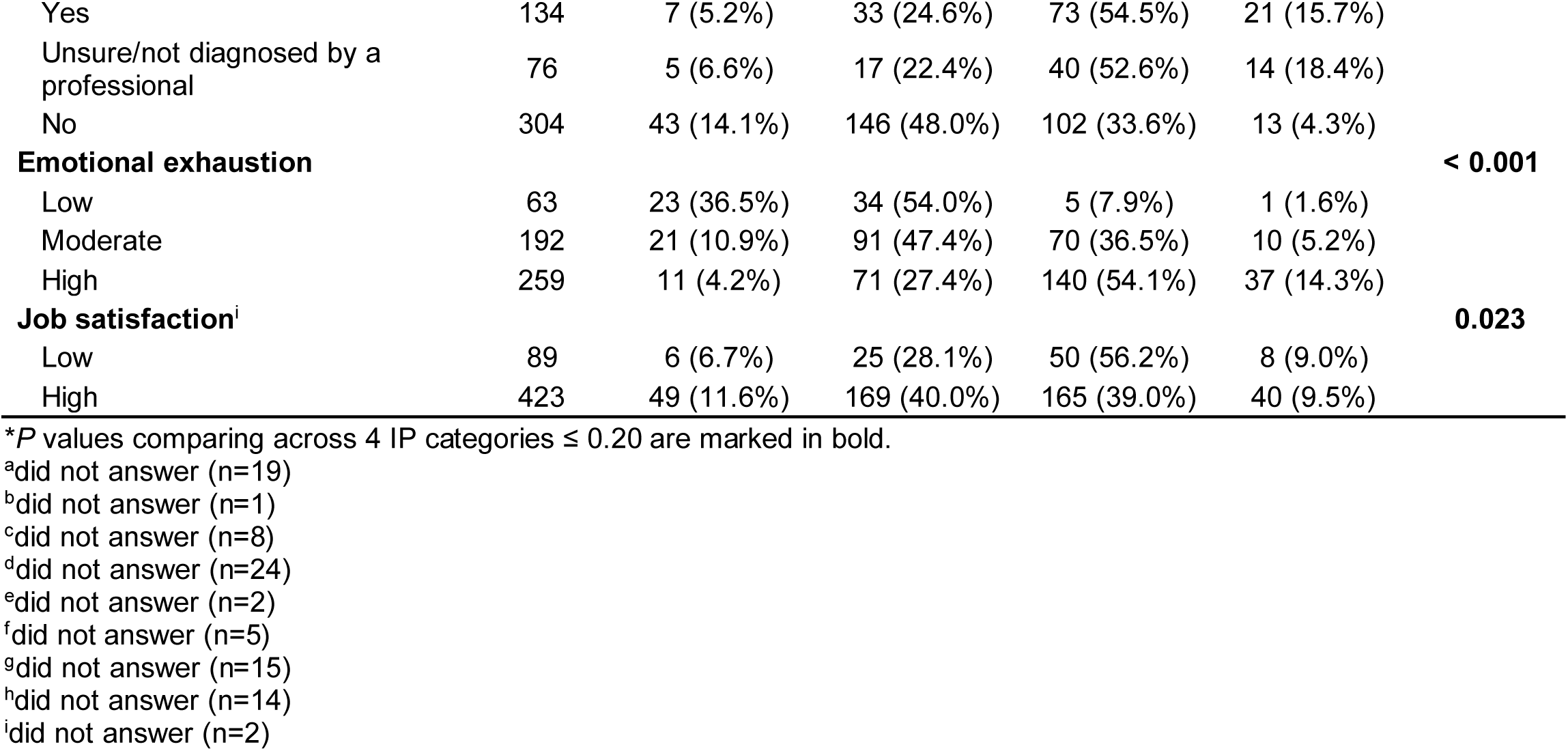
Factors Associated with IP: Multivariable Logistic Regression (n = 507)

Having managerial or supervisory experience reduced the likelihood of IP by 45% [OR=0.55, (95%CI=0.34-0.90)]. Having a terminal bachelor’s or master’s degree (versus a terminal DPT degree) [OR=2.31, (95%CI=1.07-5:00)]; history of or current mental health diagnosis [OR=2.77 (95%CI: 1.69-4.54)], unsure/undiagnosed mental health diagnosis [OR=3.21 (95%CI: 1.74-5.94)], and moderate or high level of emotional exhaustion [OR=5.37 (95%CI: 2.11-13.69); OR=14.13 (95%CI: 5.56-35.89), respectively] each increased the odds of IP. Having 11 to 20 years of clinical experience (versus ≤ 5 years) and holding their current position > 6 years (versus < 1 year) may reduce the odds of IP [OR=0.55 (95%CI: 0.28-1.08, P=0.08); OR=0.50 (95%CI: 0.24-1.04, P=0.06), respectively]. It is important to point out that these associations were cross-sectional; no causal inference could be made. For example, the negative association of having managerial or supervisory experience and the presence of IP could be bi-directional. Those with IP might not believe that they could be in a leadership role, therefore may not seek a managerial/supervisory position; or managers/supervisors were less likely to experience IP.

## DISCUSSION

IP is highly prevalent among physical therapists in the US. Over half of the survey respondents reported frequent or intense IP. Seasoned clinicians holding managerial positions exhibited a lower susceptibility to IP, whereas individuals with a mental health diagnosis, emotional exhaustion, and lacking a DPT degree were more prone to this phenomenon. Other factors, such as gender, race, sexuality, geographic location, practice specialty/setting, APTA membership, and job satisfaction did not significantly influence IP. A reduced likelihood of experiencing IP appeared to be associated with a greater length of time held in a professional position. Interestingly, there appeared to be a sweet spot in terms of years as a licensed PT. Specifically, having 11-20 years of clinical practice experience may be linked to reduced IP. To the best of our knowledge, this is the first report investigating the prevalence, predictors, and impact of IP among licensed PTs. Given the high prevalence of IP and its potential negative impacts on mental well-being, it is important to provide education and resources to effectively manage and overcome this phenomenon in the physical therapy community.

The current study findings are in general agreement with previous research conducted in other licensed health care professions. In a literature review of studies involving medical students, residents, and physicians^16^, the prevalence of IP ranged from 22.5% to 46.6%. The mean imposter scores within these studies ranged from 47.0 to 61.2.

Frequent-to-intense IP was observed in 38.9% of neurosurgeons and neurosurgery residents, with the majority of individuals reporting moderate levels of IP (42.7%)^17^. A significant proportion of radiologists (71%) experienced frequent-to-intense IP^18^. This finding should be interpreted with caution. The limited number of participants (n = 21) may introduce the possibility of participation bias, which could potentially overestimate the rate of IP.

A noteworthy result emerged from the current study, revealing that over 40% of respondents reported a diagnosed or suspected mental health condition. This finding underscores the significance of pre-existing mental health comorbidities in PTs. These conditions can be further exacerbated by elevated levels of occupational stress resulting from a number of factors, including demanding physical and mental workloads, significant administrative work (e.g., documentation and billing), high patient expectation and demands, the constant pressure to stay updated in the field and continually learn, and productivity benchmarks within health care settings^5, 19, 20^. Our data showed that respondents with diagnosed or suspected mental health conditions were approximately 3 times more likely to experience IP. Poor mental health can significantly hinder professional performance, elevate the risk of medical errors, contribute to burnout, and amplify the experience of IP^16, 21, 22^. Further, individuals with perfectionist tendencies and difficulty internalizing their successes are more susceptible to experiencing IP. These tendencies not only contribute to the development of IP but can also function as precursors to mental health conditions, such as anxiety and depression. This interplay between perfectionism, inadequate self-assessment of achievements, IP, and mental health challenges could have detrimental effects on professional and personal outcomes^16, 20, 21, 22^.

The majority of PTs (87.7%) in this survey experienced at least moderate levels of emotional exhaustion and approximately half experienced high levels. Emotional exhaustion has been identified as a central measure of burnout^23, 24^, which could lead to medical errors, reduced productivity, and turnover^5, 19^. Contrary to a previous study that reported no clear relationship between burnout and IP in 48 internal medicine residents^25^, we found a dose-response relationship between emotional exhaustion. A higher level of emotional exhaustion was associated with a greater likelihood of IP. PTs may be more susceptible to burnout, given the prolonged and frequent patient encounters within a typical work week^5^. However, in our study sample, reported job satisfaction was high among PTs (82%). Time spent on direct patient care or time spent with patients during initial evaluation and return visits did not appear to heavily impact IP. Thus, factors beyond workload and time spent with patients must be further explored to better understand the underlying causes for emotional exhaustion in PTs.

Respondents with managerial/supervisory roles or longer time in the current position were less likely to experience IP. Prior research has not consistently demonstrated a clear relationship between job ranks or professional longevity with IP. Among surgeons, IP was more prevalent and worse in trainees and residents than faculty mentors^17, 26^. On the contrary, non-tenured faculty had lower IP scores than their tenure-track counterparts^27^ and IP was common in those working in top research institutions^28^. This is potentially due to the high-pressure research environments and intense competition for funding and recognition. The conflicting observations could be related to differences in job security, workload, and expectations between clinical and academic settings.

Gender was not linked to IP when accounting for other demographic, professional, and mental health factors. While early studies primarily focused on the prevalence of IP in females in various occupational settings, updated research revealed that IP is common in both men and women^1^. Notably, the univariable association between gender and IP attenuated to non-significance after adjusting for other variables. This finding highlights that gender alone may not be a significant predictor of IP and caution against drawing conclusions solely on gender when seeking to understand the risk profile of IP.

The present study has significant strengths. This is the first study that comprehensively examined IP and its predictors among licensed PTs, using a validated outcome measure to categorize IP. Compared to previous research of IP in health care professionals, the present study included the largest sample to date and employed a more rigorous analytic approach of multivariable regression modeling. Our study sample was representative of the demographics of licensed PTs in the United States, as reported by the U.S. Bureau of Labor Statistics, 2022^29^. This alignment with the broader PT community enhances the generalizability of our findings and allows for meaningful comparisons and inferences regarding IP in PTs. Additionally, a focus group was utilized prior to survey distribution to ensure inclusivity and ease with the survey experience.

### Limitations

Our study also has limitations. The cross-sectional study design precludes us from establishing causal relationships. We cannot definitively determine whether the identified demographic and professional factors were predictors for or consequences of IP. Nonetheless, this study provides a valuable groundwork for future investigations aimed at mitigating potential risk factors associated with IP. Although this survey was peer reviewed by a focus group of PTs from diverse clinical settings to ensure clarity and comprehensibility prior to its dissemination, some variability in the interpretation of the survey questions may have occurred. Due to the multiple channels of survey distribution and de-identified collection of information, individuals may have accessed and responded to the survey more than once, despite being informed to avoid repeated attempts.

### Conclusions

IP is highly prevalent among licensed PTs, with more than half reporting frequent or intense IP. Seasoned clinicians with managerial roles seemed to be less susceptible to IP, whereas those with mental health diagnoses, emotional exhaustion, and those without a DPT degree may be more likely to experience IP. Given the high prevalence of IP and its negative impacts, it is important to prioritize education and awareness about this phenomenon. PT clinicians and students need to be equipped with information, strategies, and resources to effectively manage and mitigate IP, ensuring their long-term professional success and personal well-being.

## Supporting information

STROBE Checklist

## Data Availability

All data produced in the present study are contained within the manuscript.

---

**Supplemental TABLE 1.**
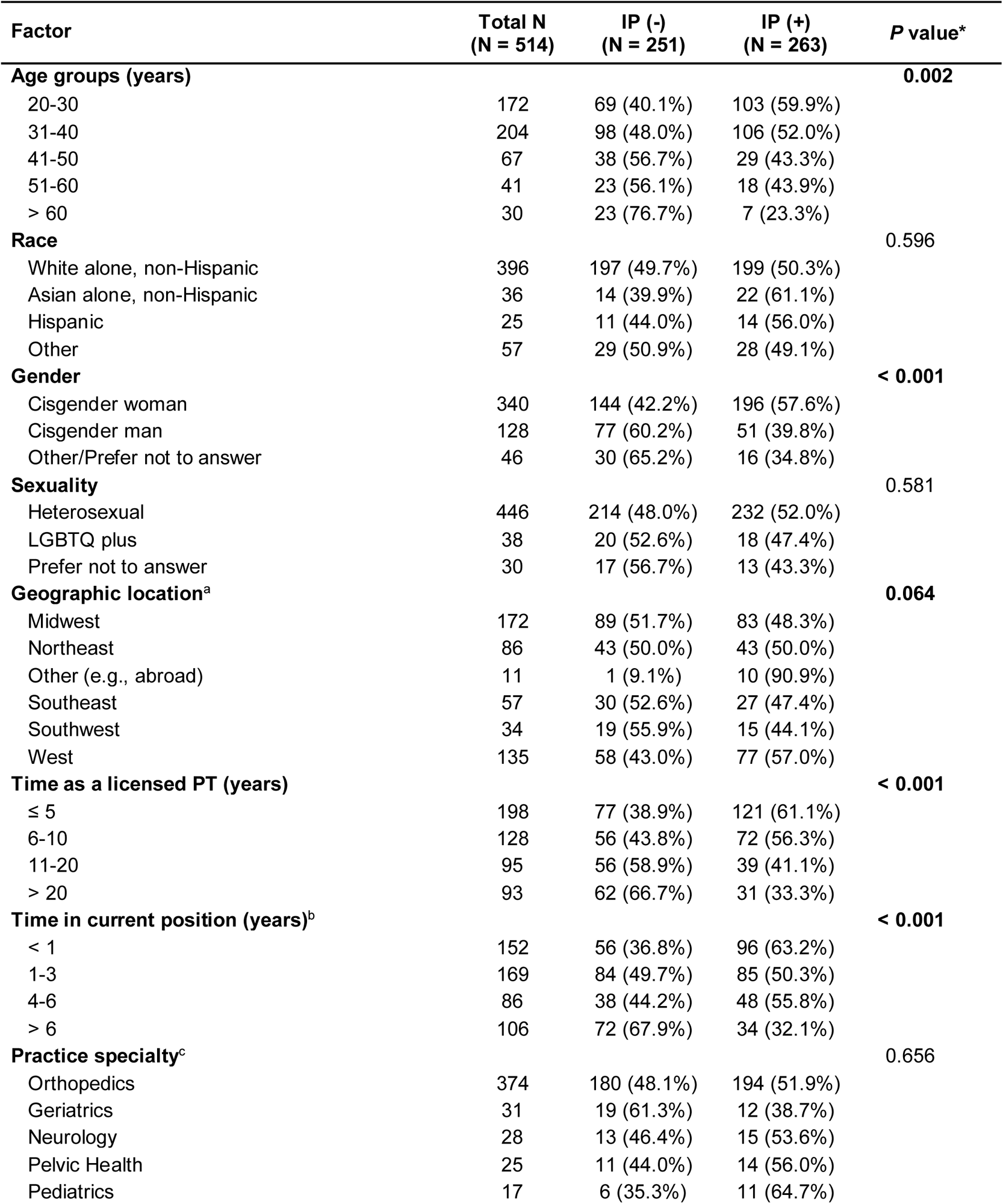
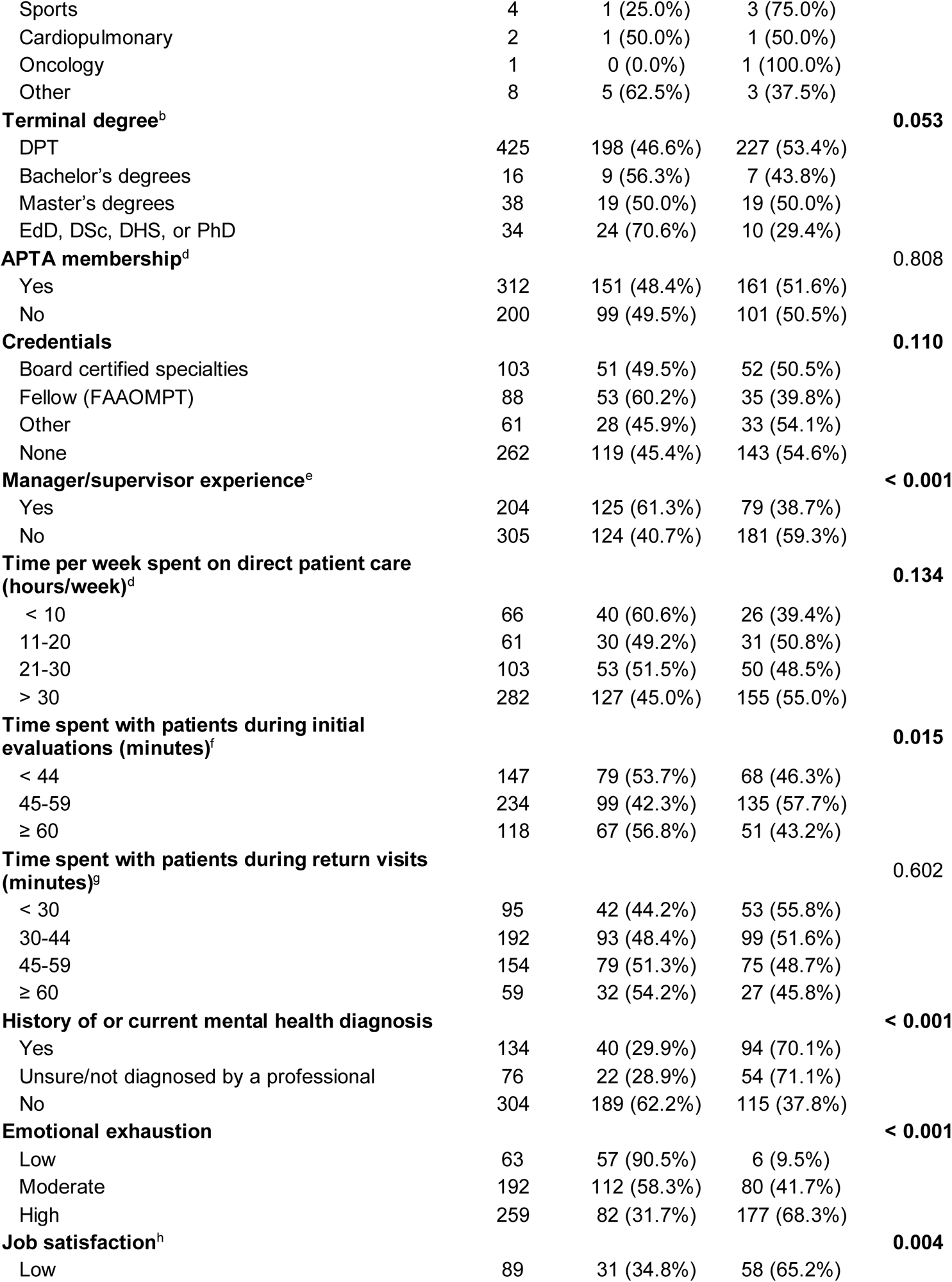
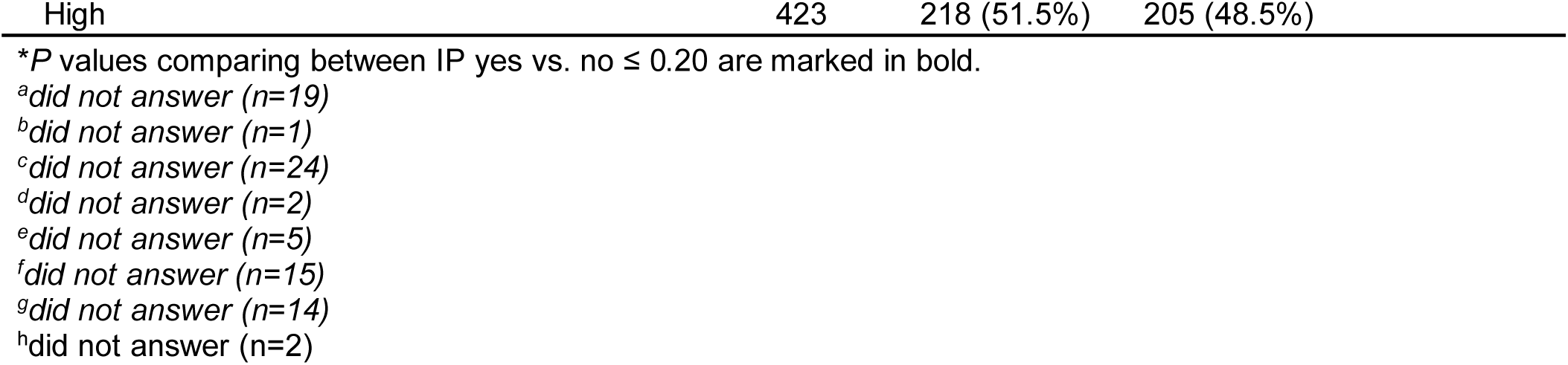
Descriptive Summary of Demographic and Professional Factors related to IP (yes vs. no) (Univariate Comparisons)

## Acknowledgements

The authors would like to acknowledge and thank the Orthopedics, Geriatrics, Neurology, and Home Health academies of the American Physical Therapy Association and the American Academy of Orthopaedic Manual Physical Therapists for their assistance and contribution to this study.

